# Exploring the extracellular transcriptome in seminal plasma for non-invasive prostate cancer diagnosis

**DOI:** 10.1101/2021.05.11.21256306

**Authors:** Eva Hulstaert, Annelien Morlion, Justine Nuytens, Giovanni Ponti, Monia Maccaferri, Susan Lau, Eleftherios Diamandis, Jarvi Keith, Ioannis Prassas, Nicolaas Lumen, Pieter Mestdagh, Jo Vandesompele

## Abstract

A diagnostic non-invasive biomarker test for prostate cancer at an early stage, with high sensitivity and specificity, would improve diagnostic decision making. Extracellular RNAs present in seminal plasma might contain biomarker potential for the accurate detection of clinically significant prostate cancer. So far, the extracellular messenger RNA (mRNA) profile of seminal plasma has not been interrogated for its biomarker potential in the context of prostate cancer. Here, we investigate the mRNA transcriptome in seminal plasma samples obtained from prostate cancer patients (n=25), patients with benign prostate hyperplasia (n=26) and individuals without prostatic disease (n=6). Seminal plasma harbors a complex mRNA repertoire that reflects prostate as its tissue of origin. The endogenous RNA content is higher in the prostate cancer samples compared to the control samples. Prostate cancer antigen 3 (PCA3), a long non-coding RNA with prostate cancer-specific overexpression, and ATP-binding cassette transporter 1 (ABCA1), known to be involved in the prostate cancer pathogenesis, were more abundant in the prostate cancer group. In addition, twelve high confidence fusion transcripts could be detected in prostate cancer samples, including the bona-fide prostate cancer fusion transcript TMPRSS2-ERG. Our findings provide proof-of-principle that the extracellular transcriptome of seminal plasma can reveal information of an underlying prostate cancer.

## Introduction

Prostate cancer is the most common internal cancer in men and an important cause of cancer death in developed countries^1^. Prostate cancer is mostly diagnosed at a localized stage through a combination of serum prostate-specific antigen (PSA), digital rectal examination, magnetic resonance imaging and prostate biopsy^2^.

The introduction of serum PSA testing has revolutionized the prostate cancer diagnostics field. However, this biomarker has a number of limitations. Although PSA is specific for prostatic tissue, this marker has low prostate cancer specificity, as PSA is elevated in other nonmalignant pathologies of the prostate, including benign prostate hyperplasia (BPH) and prostatitis. More than half of the patients presenting with an elevated PSA level have a negative prostate biopsy result^3^. In addition, PSA is not able to differentiate between clinically significant (and thus potentially lethal) and insignificant prostate cancer. PSA screening thus leads to many unnecessary biopsies of benign disease and diagnosis of clinically non-significant and non-evolving cases of prostate cancer.

There is a clinical need for robust, non-invasive biomarkers that accurately detect prostate cancer and differentiate indolent from life-threatening disease. The ideal test would be minimally invasive, have few to no side effects, identify a high proportion of men who would benefit from treatment, and minimize the identification of men with clinically insignificant cancer in order to prevent overtreatment.

In an attempt to identify biomarkers, genomic, transcriptomic, epigenetic and proteomic methods are applied to profile prostate cancer tissues and biofluids obtained from prostate cancer patients. RNA sequencing allows the profiling of the extracellular transcriptome in human biofluids, including seminal plasma. We recently demonstrated that the mRNA concentration in seminal plasma is about 1000-fold higher than the RNA concentration in platelet-free plasma or urine, the most studied biofluids in the context of prostate cancer screening^4^. Seminal plasma also contains a richer mRNA content of about 11 000 unique mRNAs that can be detected in 200 µL seminal plasma, while only 2000 unique mRNAs are detected in 200 µL urine or platelet-free plasma, suggesting that seminal plasma has a high biomarker potential. Prostate cancer antigen 3 (PCA3), a long non-coding RNA with prostate cancer-specific overexpression, is the most widely studied RNA biomarker in this field. The Progensa PCA3 urine test measures the concentration of PCA3 and PSA RNA and calculates the ratio of PCA3 RNA to PSA RNA in post-digital rectal exam first-catch urine specimens. The test was approved by the American Food and Drug administration (FDA) for helping clinicians whether or not to recommend a repeat biopsy after a negative biopsy result ^5^. However, the use of PCA3 testing is not recommended in clinical setting due to the limited evidence that it supports improved clinical outcome^6^. From a biological point of view, seminal plasma holds great potential for biomarker discovery as it shows proximity to the prostate tumor and it can be collected in a noninvasive way^7–9^. Approximately 40% of semen is derived from prostatic tissue^10^ and prostate specific mRNA signatures are enriched in RNA sequencing data of seminal plasma compared to other biofluids^4^. Of note, PSA was originally discovered in seminal plasma, where its concentration is about 5-6 times higher than in serum^11^. Small RNA sequencing has been performed on the non-sperm cellular fraction of seminal fluid (including prostatic epithelial, urothelial and inflammatory cells) on two pools representing 6 men with prostate cancer and 6 men without cancer. A higher ratio of transfer RNA (tRNA) to microRNA (miRNA) was observed in the cancer pool compared to the control pool.^12^ The biomarker potential of specific miRNAs in cells obtained from semen samples and in extracellular vesicles isolated from semen has also been assessed using reverse transcription quantitative polymerase chain reaction (RT-qPCR) ^13, 10^. The full mRNA content of seminal plasma obtained from prostate cancer patients has not been studied yet.

The main goal of this study is to investigate the biomarker potential of extracellular RNA in seminal plasma as a non-invasive approach for prostate cancer diagnosis. Here, we present mRNA capture sequencing data of 57 seminal plasma samples from 25 prostate cancer patients, 26 BPH patients and 6 healthy controls.

## Material and methods

### Donor material, collection and seminal plasma preparation procedure

Seminal plasma was collected in prostate cancer patients, in patients with benign prostate hyperplasia and in healthy donors. The diagnosis of prostate cancer was histologically proven on prostate biopsy and/or on radical prostatectomy specimen. For all patients with benign prostate hyperplasia there was no evidence of prostate cancer on prostate biopsy. All samples were collected prior to prostate biopsy or prostatectomy. A prostate biopsy was not performed in the healthy donors. The study samples were collected at three urologic centers. Sample collection was approved by the ethics committee of Ghent University Hospital, Belgium (no. B670201734450), the ethics committee of the University of Modena and Reggio Emilia, Italy, and the Murray Koffler Urologic Wellness Centre at Mount Sinai Hospital, Canada (no. 08-0117-E). Written informed consent was obtained from all donors according to the Helsinki declaration.

Semen samples were produced by masturbation and collected into a sterile container. Samples collected at Ghent University Hospital were allowed to liquefy for 30 min at 37 °C. Samples were centrifuged to remove cells (2000 g, 10 min) and stored at -80 °C within 2 hours after collection. Samples collected at the University of Modena and Reggio Emilia were allowed to liquefy for a maximum of 2 h at room temperature. Samples were prepared with a 2-spin protocol at room temperature: the samples were first centrifuged at low speed (400 g, 10 min), the supernatant was then removed from the cell pellet. Next, the samples were centrifuged at high speed (16,000 g, 5 min) and the supernatant was stored at -80 °C. Samples collected at Mount Sinai Hospital were allowed to liquefy for 1 h at room temperature. Samples were centrifuged at high speed (13,000 g, 15 min) and the supernatant was stored at -80 °C.

### RNA isolation and gDNA removal

Total RNA was purified with the miRNeasy Serum/Plasma Kit (Qiagen, Hilden, Germany, #217184), starting from an input volume of 200 µL, according to the manufacturer’s instructions. Per 200 µL seminal plasma input volume, 2 µL Sequin spike-in controls (1/1300 of the stock solution, Garvan Institute of Medical Research, mix A) were added prior to the RNA purification. After the RNA purification, 2 µl External RNA Control Consortium (ERCC) spike-in controls (1/1000 of the stock solution, ThermoFisher Scientific, Waltham, MA, USA, #4456740, mix 1), 1 µl HL-dsDNase and 1.6 µl reaction buffer were added to 12 µl RNA eluate, and incubated for 10 min at 37 °C, followed by 5 min at 55 °C to remove contaminating genomic DNA. An overview of the Sequin and ERCC spike-in controls with their stock concentration is provided in supplemental table 1.

### TruSeq RNA Exome library prep sequencing

Messenger RNA capture based libraries were prepared starting from 8.5 µL DNase treated RNA eluate using the TruSeq RNA Exome Library Prep Kit (Illumina, San Diego, CA, USA, #20020189, #20020490, #20020183), as previously described^14^. This library preparation protocol enriches for exonic RNA sequences. Each sample underwent individual enrichment according to the manufacturer’s protocol. The quality and yield of the prepared libraries were assessed using a high sensitivity Small DNA Fragment Analysis Kit (Agilent Technologies, Santa Clara, CA, USA, #DNF-477-0500) according to manufacturer’s instructions. Only samples with a fragment analyzer profile of a good quality cDNA library (i.e. a band around 260 bp before capture) were included for further analysis. The libraries were quantified using qPCR with the KAPA Library Quantification Kit (Roche Diagnostics, Diegem, Belgium, #KK4854) according to manufacturer’s instructions. Based on the qPCR results, equimolar library pools were prepared.

Paired-end sequencing was performed on a NextSeq 500 instrument using a high output v2 kit (Illumina, San Diego, CA, USA) with a read length of 75 nucleotides. All samples were sequenced twice with a loading concentration of 2 pM (2% PhiX). The FASTQ files of both runs were combined, resulting in an average sequencing depth of 9 million read pairs per sample.

### Data analysis

#### Processing TruSeq RNA Exome sequencing data

Read quality was assessed by running FastQC (v0.11.5) on the FASTQ files and reads shorter than 35 nucleotides and with a quality (phred) score < 30 were removed. The reads were mapped with STAR (v2.6.0). Clumpify (v38.26) was used for duplicate read removal. HTSeq (v0.9.1) was used for quantification of deduplicated reads. Mapped reads were annotated by matching genomic coordinates of each read with genomic locations of mRNAs (obtained from UCSC GRCh38/hg38 and Ensembl, v91). Reads were subsampled after duplicate removal to 500,000 reads per sample. Only samples with a total Sequin spike coverage of at least 100 reads were retained. A cut-off of 2 counts per gene was applied to remove low abundant genes. The mass of endogenous mRNA present in 1 mL seminal plasma was estimated based on the read count for ERCC-00130 spike-in RNA. To each RNA eluate sample, 6E-17 mol of ERCC-00130 was added (molecular weight of 340415.55 g/mol). Based on the read count for the endogenous mRNA, the corresponding mass of endogenous mRNA in the eluate was calculated and subsequently corrected for input volume.

#### Assessment of tissue and cell contribution to the extracellular transcriptome of seminal plasma

Using total RNA-sequencing data from 27 normal human tissue types and 5 immune cell types from peripheral blood from the RNA Atlas^15^, we created gene sets containing marker genes for each individual entity as described^4^. We removed redundant tissues and cell types from the original RNA Atlas (e.g. granulocytes and monocytes were present twice; brain was kept and specific brain sub-regions such as cerebellum, frontal cortex, occipital cortex and parietal cortex were removed) and we used genes where at least one tissue or cell type had expression values greater or equal to 1 TPM normalized counts. A gene was considered to be a marker if its abundance was at least 5 times higher in the most abundant sample compared to the others. For the final analysis, only tissues and cell types with at least 3 markers were included, resulting in 26 tissues and 5 immune cell types. Gene abundance read counts from the biofluids in the discovery cohort of Hulstaert et al. and the gene abundance read counts of the seminal plasma cohort from this study, were normalized using Sequin spikes as size factors in DESeq2 (v1.22.2)^4^. For all marker genes within each gene set, we computed the log2 fold changes between the median read count of the seminal plasma samples versus the median read count of all other biofluids.

#### Differential expression analysis with DESeq2

Further processing of the count tables was done with R (v3.5.1) making use of tidyverse (v1.2.1). Gene abundance expression read counts from the seminal plasma samples were normalized using the sum of all reads mapping to Sequin spikes as size factors in DESeq2 (v1.20.0)^16^. The Sequin normalized count table is provided in supplemental table 3. To assess the biological signal in seminal plasma, we performed differential expression analysis between the patients and control groups using DESeq2 (v1.20.0). Genes were considered differentially expressed when the absolute log2 fold change > 1 and q < 0.05.

#### Differential exon usage with DEXSeq

To perform differential exon usage analysis the mapped sequencing data was preprocessed according to the two preparation Python scripts provided in the DEXSeq package (version 1.36.0, ^17^). In the first script, a GTF file with gene models was transformed into a GFF file listing counting bins. In the second script such a GFF file and an alignment file in the BAM format were used to produce a list of exon counts. Next, the count tables consisting of exon counts were further processed with R (v3.5.1) making use of tidyverse (v1.2.1). Exon expression read counts were normalized using the sum of all reads mapping to Sequin spikes as size factors in DESeq2 (v1.20.0)^16^. Differential expression analysis between the patients and control groups was performed using DESeq2 (v1.20.0). Exons were considered differentially expressed when the absolute log2 fold change > 1 and q < 0.05. In order to identify genuine differentially abundant exons, only exons that were not part of differentially abundant genes were considered.

#### Gene set enrichment analysis

A preranked gene set enrichment analysis was performed using the hallmark gene sets (v7.2) and the Kyoto Encyclopedia of Genes and Genomes (KEGG) canonical pathways of the curated gene sets (v7.2) available on the Molecular Signatures Database, applying 1000 permutations and a classic enrichment statistic^18^. All mRNA lists were ordered based on the log-transformed fold change obtained after differential expression analysis with DESeq2. Gene sets with a false discovery rate (FDR) up to 0.25 were selected for exploratory discovery of candidate hypotheses.

#### Detection of fusion transcripts

Fusion transcript identification was performed using FusionCatcher (v1.30) with default parameter settings^19^. Stringent filtering was applied to exclude potential false positive fusion transcripts, as previously described^20^. First, transcripts with a fusion description label indicative for a false positive result (i.e. the red annotations in supplemental table 5) were excluded. Second, transcripts with reads mapping on both fusion partners were excluded. Third, transcripts with fusion partners less than 100 kbp apart were also excluded. Only exon-exon fusions were included.

## Results

### Patient population

Only 57 of the 104 seminal plasma samples that were collected for this study met the inclusion criteria described in the materials and methods section and were included for further analysis in this study (Figure 1). In 17 samples, no eluate was obtained at the end of the RNA purification protocol, probably due to the high viscosity of the samples leading to a clogged RNA purification column. In 6 samples, no cDNA library was present on fragment analyzer during the mRNA capture library preparation protocol, indicating that the RNA was too degraded or too low concentrated in these samples. In the remaining samples, the sequencing depth varied from 7 million to 32 million reads with a median of 15 million reads per sample. The percentage of PCR duplicates ranged from 51% to 99% with a median of 90%. High PCR duplicate levels are expected in samples with low RNA concentration. After the removal of reads with low quality, after adapter trimming and after the removal of PCR duplicates, the remaining reads per sample ranged from 122,650 to 8 million with a median of 1 million reads per sample. A minimum threshold of 500,000 reads per sample was applied, resulting in the exclusion of another 25 samples. One sample was excluded because of a low coverage of Sequin spikes (fewer than 100 reads), not allowing a robust normalization using Sequin spikes. An overview of the reads obtained per sample and per step of the pre-processing workflow are provided in supplemental table 2.

**Figure 1.**
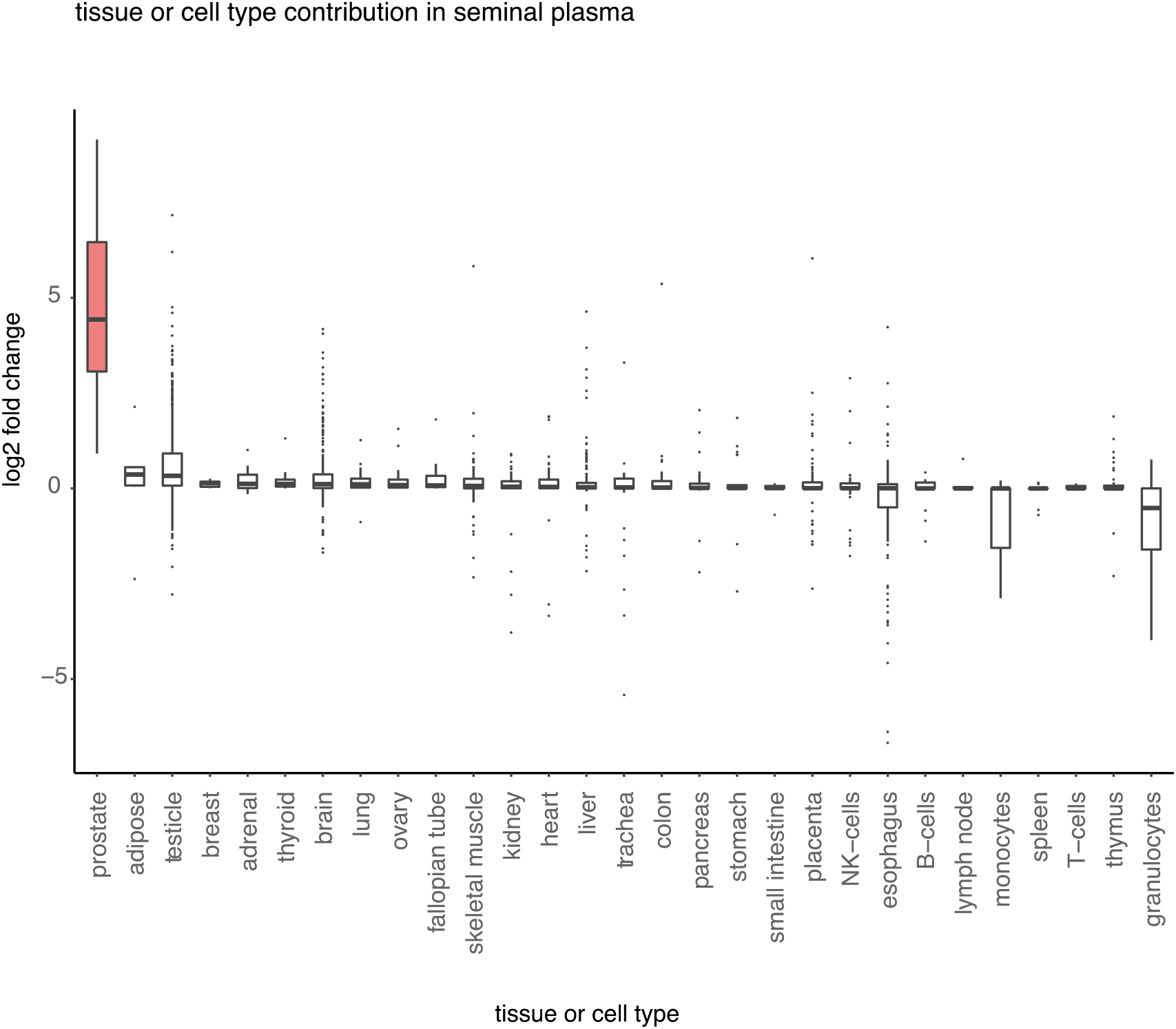
(A) Assessment of normal human tissues and cell types contributing to seminal plasma extracellular RNA. Boxplots showing the log_2_ fold change for a gene set with markers specific for a certain tissue or cell type. The log_2_ fold change is calculated between the median read count of all seminal plasma samples and the median read count of all other biofluids. The tissues or cell types for which markers were selected based on the RNA Atlas Project are shown on the x-axis.

The statistical analyses were thus performed on 57 seminal plasma samples obtained from 25 prostate cancer patients with a mean age of 65 years old, 26 patients with benign prostate hyperplasia with a mean age of 60 years old and 6 healthy individuals with a mean age of 60 years old. Most prostate cancer patients (18/25) showed well to moderately differentiated disease (Gleason score 6 or 7). The average PSA level in the prostate cancer group and in the BPH group was 9.08 ng/mL and 4.90 ng/mL, respectively. Demographics and clinical details for the included samples are shown in Table 1. Details for all samples with the reason of exclusion for further analysis is provided in supplemental table 2. Principal component analysis of all detected genes did not reveal clustering based on the collection site, indicating that the seminal plasma preparation protocol that slightly differs between the three collection sites is not a dominant driver of mRNA abundance variance in our cohort. In addition, this analysis did not reveal clustering of samples according to the clinical diagnosis of the donor (supplemental figure 2).

**Table 1.** Clinical details of patient samples included for further analysis

|  | cancer (n=25) | BPH (n=26) | healthy individual (n=6) |
| --- | --- | --- | --- |
| age in years, mean (min-max) | 65 (46-78) | 60 (45-69) | 60 (56-66) |
| pre-operative PSA in ng/mL, mean (min-max) | 9.08 (3.00-31.00) | 4.90 (0.70-10.00) | - |
| Gleason score |  |  |  |
| 6 | 9 | - | - |
| 7 (3+4) | 6 | - | - |
| 7 (4+3) | 3 | - | - |
| 8 | 6 | - | - |
| 9 | 1 | - | - |
| collection center |  |  |  |
| Belgium | 6 | 0 | 0 |
| Canada | 14 | 14 | 0 |
| Italy | 5 | 12 | 6 |

### Seminal plasma harbors a complex mRNA repertoire containing prostate specific mRNAs

The mapping rate varied from 67% to 99% with a median mapping rate of 98% (supplemental figure 3A). The total number of unique mRNAs detected with at least 2 counts per sample ranged from 7050 to 12,464 with a mean of 10,089 mRNAs per sample (supplemental figure 3A). In total, 2418 genes were detected in all samples. Prostate specific mRNAs are most abundant in seminal plasma compared to other tissue specific signatures, supporting the exploration of the biomarker potential for prostate cancer detection in this fluid (figure 1). The latter finding is reflected at individual mRNA level. Prostate specific mRNAs such as KLK3 (kallikrein related peptidase 3 or prostate specific antigen, PSA) and TGM4 (transglutaminase 4) are highly abundant in all samples, while liver specific mRNAs such as FMO3 (flavin containing monooxygenase 3) and CYP2C8 (cytochrome P450 family 2 subfamily C member 8) are detected in none of the samples (supplemental table 3).

Based on the coverage of artificial spike-in controls, the endogenous RNA mass per sample was calculated. The mean endogenous RNA mass detected per 1 mL fluid was 86.13 ng in the prostate cancer group (minimum 0.64 ng, maximum 307.75 ng) and 51.20 ng in the control group (minimum 0.33 ng, maximum 686.52 ng). The endogenous RNA mass was higher in the prostate cancer group compared to the control group (Wilcoxon signed-rank test, two-sided, p = 0.0429, supplemental figure 3B).

### PCA3 and ABCA1 are higher in cancer samples

Differential expression analysis revealed 2 mRNAs that are more abundant in seminal plasma from prostate cancer patients compared to controls, consisting of patients with BPH and healthy individuals (Figure 4A). A list with the results of the differential expression analysis can be found in supplemental table 3. The normalized counts of PCA3 and ABCA1 in both groups are shown in figure 4B. Gene set enrichment analysis for the contrast comparing prostate cancer versus controls, consisting of patients with BPH and healthy controls, demonstrated an enrichment of genes that are involved in spermatogenesis, coagulation, inflammatory responses and KRAS signaling, when looking into the hallmark gene sets (figure 4C). Investigation of the KEGG canonical pathways of the curated gene sets points towards an enrichment of genes involved in lysosomal activity (figure 4C). Off note, lysosomal enzymes have been hypothesized to be involved in the pathogenesis of prostate cancer^21,22^. In all three contrasts (prostate cancer versus all controls; prostate cancer versus patients with BPH; prostate cancer versus healthy individuals), a consistent lower abundance of genes involved in the androgen response and the oxidative phosphorylation and MYC target genes was observed.

**Figure 3.**
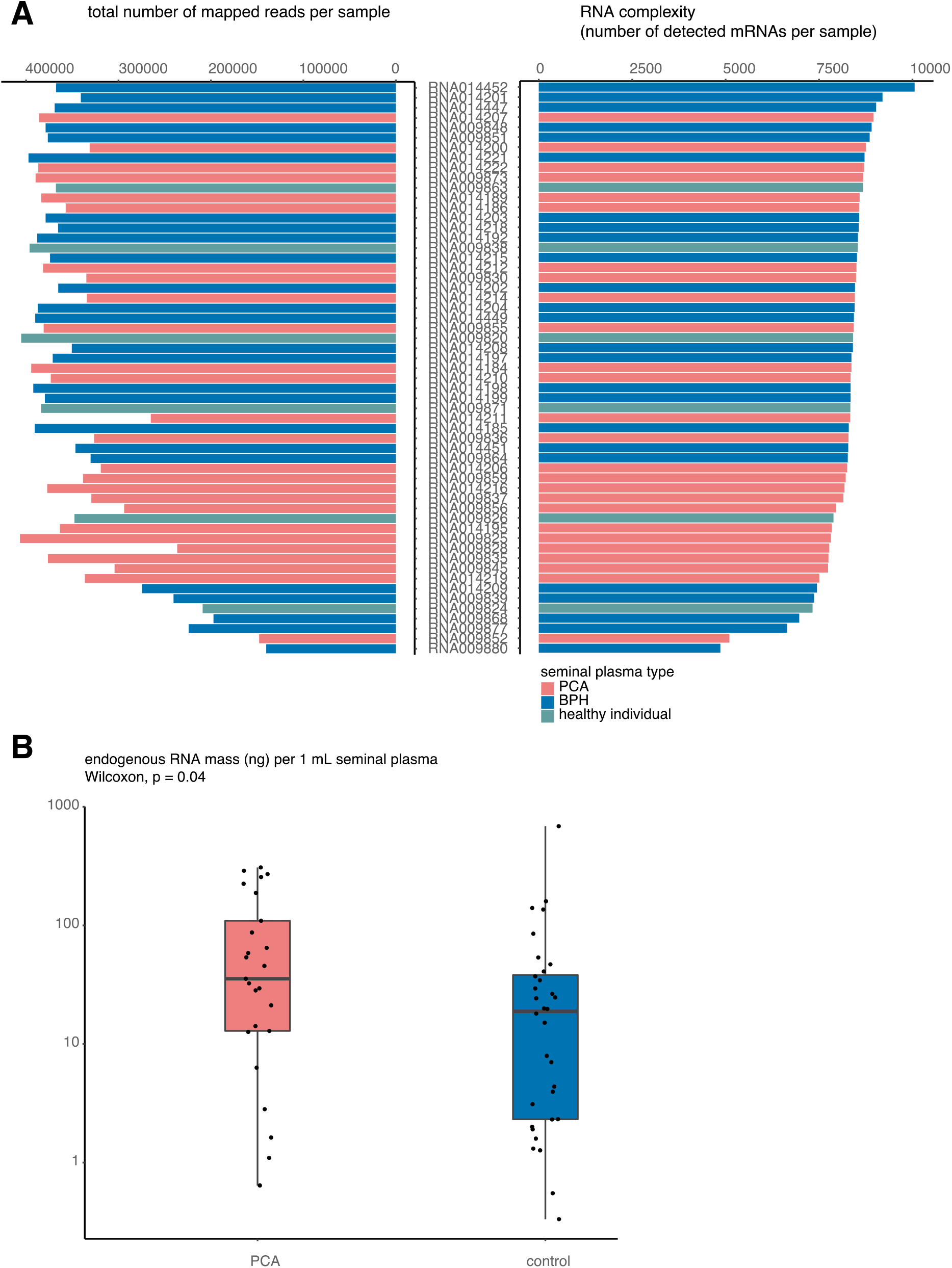
(A) Barplots showing the percentage of the total read count mapping to the human transcriptome per sample (left) and the total number of unique mRNAs that are detected per sample (right). (B) The endogenous RNA mass in ng detected per 1 mL fluid compared between the control samples (n=32; consisting of 26 samples from patients with benign prostate hyperplasia and 6 samples from healthy individuals) and the prostate cancer samples (n=25; Wilcoxon-signed rank test, two-sided, p=0.0429). PCA, prostate cancer; BPH, benign prostate hyperplasia

**Figure 4.**
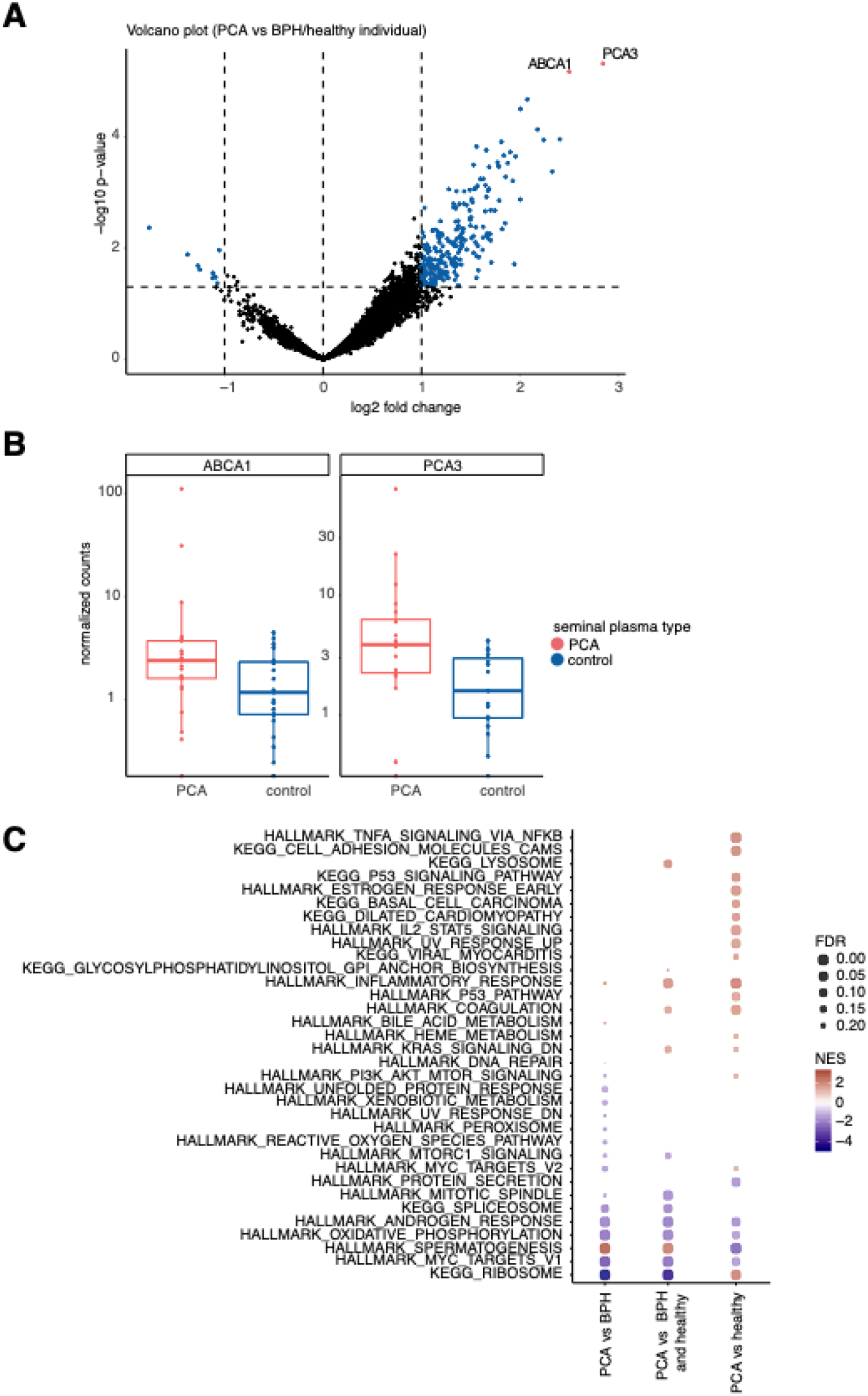
(A) Volcano plot of differentially abundant mRNAs in prostate cancer versus controls (consisting of benign prostate hyperplasia samples and samples obtained in healthy individuals). Up- and downregulated genes with a p-value of less than 0.05 are shown in blue. Upregulated genes with an adjusted p-value of less than 0.05 are shown in pink. (B) Boxplots comparing the Sequin spike normalized read counts per group. The normalized read count per sample is shown as a dot. Samples obtained from prostate cancer patients are pink, samples obtained from patients with benign prostate hyperplasia or from healthy volunteers are blue. (C) Gene set enrichment analysis results for the hallmark gene sets and KEGG canonical pathways of the curated gene sets. Normalized enrichment score (NES) and false discovery rate (FDR) are depicted on the enrichment plot. All gene sets with a false discovery rate (FDR) < 0.25 are shown. BPH, benign prostate hyperplasia; NES, normalized enrichment score; PCA, prostate cancer; pval, p-value; padj, adjusted p-value.

When applying the differential expression analysis on the prostate cancer and BPH samples only, no differentially expressed genes remained after multiple testing correction (data not shown). The differential expression analysis was also performed on exon level instead of gene level; also here no differential exons were identified in this dataset.

### Bona-fide prostate cancer fusion transcripts are detected in cancer samples

In total 923 fusion transcripts were detected in 20 prostate cancer samples and 26 control samples. After stringent filtering, 12 high confidence fusion transcripts remained in 11 prostate cancer samples and 2 high confidence fusion transcripts remained in 4 control samples. An overview of the high confidence fusion transcripts is provided in table 2. The detailed output of FusionCatcher is provided in supplemental table 5. Amongst the detected fusion transcripts in prostate cancer samples was TMPRSS2-ERG (transmembrane protease, serine 2 - v-ets erythroblastosis virus E26 oncogene homolog), a fusion gene that is reported to be present in approximately 50% of prostate cancer lesions^23,24^. Another detected fusion transcript in one prostate cancer sample was TMPRSS2-BRAF (transmembrane protease, serine 2 - v-raf murine sarcoma viral oncogene homolog B1). While this transcript is not frequently reported in prostate cancer, it was observed in one prostate cancer tissue sample when consulting the TumorFusions data portal. This portal contains an overview of all fusion transcripts detected in 9966 cancer samples and 648 normal specimens from The Cancer Genome Atlas ^25^. The remaining 10 fusion transcripts have not been reported yet.

**Table 2.** High confidence fusion transcripts detected in seminal plasma

| gene fusion | fusion point of the 5' end fusion partner | fusion point of the 3' end fusion partner | seminal plasma sample | group |
| --- | --- | --- | --- | --- |
| TMPRSS2-ERG | 21:41498119:- | 21:38445621:- | RNA014221 | cancer |
| TMPRSS2-BRAF | 21:41488394:- | 7:140781693:- | RNA009851 | cancer |
| SPIN1-ST13 | 9:88416766:+ | 22:40848369:- | RNA014204 | cancer |
| DIO2-CEP128 | 14:80387678:- | 14:80530886:- | RNA014452 | cancer |
| PARG-BMS1 | 10:49885203:- | 10:42791627:+ | RNA014452 | cancer |
| BMS1-PTPN20 | 10:42787247:+ | 10:46984230:+ | RNA014452, RNA014447 | cancer |
|  | 10:42793991:+ | 10:46984230:+ | RNA009820 | healthy individual |
| C9ORF85-C19ORF47 | 9:71947112:+ | 19:40332643:- | RNA014192 | cancer |
| GRIPAP1-CBFA2T2 | X:49000566:- | 20:33606956:+ | RNA014192 | cancer |
| CACNB2-ABCC4 | 10:18522238:+ | 13:95083290:- | RNA014201 | cancer |
| RPS24-LIAS | 10:78040696:+ | 4:39468874:+ | RNA014204 | cancer |
| AC022400.4-PARG | 10:73713204:- | 10:49869555:- | RNA009848, RNA014449 | cancer |
| FCN1-PTMS | 9:134905890:- | 12:6770157:+ | RNA009835, RNA009845, RNA009855 | BPH |

## Discussion and future perspectives

Analysis of seminal plasma is particularly interesting to prostate diseases, such as prostate cancer, because of its proximity to the prostate and its noninvasive collection method. Only a few studies have looked into the content of seminal plasma. Previous studies have focused on cell-free DNA^8^ and the proteome ^7,26,27^ for biomarker discovery. To our knowledge, this is the first time that the full repertoire of mRNAs has been successfully studied in seminal plasma samples from prostate cancer patients.

A major limitation of our study is the substantial proportion (45%, 47 out of 104) of collected samples that did not meet the minimum quality criteria for mRNA capture sequencing. Potential explanations for the high number of excluded samples can be differences in pre-analytical variables. Liquefaction time and centrifugation speed to prepare seminal plasma was different in the three participating centers. To overcome the differences in pre-analytical variables, a dedicated prospective sample collection with a standardized protocol is important. In 17/104 samples no RNA eluate was obtained in the last step of the RNA purification, suggesting that the RNA purification column got clogged, probably due to the viscosity of the seminal plasma. This also indicates that the RNA isolation method for this specific biofluid needs further optimization. A bead-based RNA purification method may be better suited for this biofluid compared to a column-based isolation protocol. The inclusion rate of 55% (57/104) in our study, is in line with the reported inclusion rate of 43% (66/152)^13^. In the latter study, total RNA was isolated from the epithelial cell layer of semen samples and specific microRNAs were investigated using RT-qPCR. The RNA diversity (reported as the unique number of detected mRNAs per sample) observed in our study is in line with the previously reported number of 11,868 unique mRNAs in 200 µL seminal plasma of healthy donors^4^. Note that no read downsampling was applied in the Human Biofluid Atlas, resulting in a slightly higher RNA diversity.

Although the RNA diversity, and thus the theoretical chance to identify new biomarkers, in seminal plasma is higher than in urine or blood-derived plasma, we could only detect two statistically significant, differentially abundant mRNAs in this study. PCA3 and ABCA1 are significantly more abundant in seminal plasma from prostate cancer samples compared to controls (consisting of benign prostate hyperplasia samples and samples from healthy volunteers). PCA3 has been extensively studied as biomarker for prostate cancer in urine collected after prostatic massage or digital rectal examination. ABCA1 has previously been linked to the pathogenesis of prostate cancer^28–31^. ABCA1 promotor hypermethylation and subsequent transcriptional silencing has been described as a mechanism used by prostate cancer cells to maintain elevated intracellular cholesterol levels. Intracellular cholesterol has two proposed roles in the development of advanced prostate cancer: serving as a substrate in de novo androgen synthesis in castration-resistant prostate cancer and enhancing AKT signaling by stabilizing lipid raft structure^29^.

Bulk RNA sequencing allows inspection of the entire spectrum of RNA in a biofluid, including tumor-derived signals as well as signals from the tumor-microenvironment. The majority of the RNA signals obtained in this study are most likely derived from non-malignant (prostate and testicular) cells. The fraction of RNA derived from prostatic cancer cells in our cohort containing mainly Gleason 6 and 7 tumors, may be too low to obtain a clear tumor-derived signal. Most of the cancer samples included in this cohort are indeed obtained in patients with well- to moderately differentiated disease. This might also explain the relatively low number of differentially abundant mRNAs detected in this study. The isolation of prostate cancer cells from seminal plasma and the isolation of cellular RNA might be an alternative, though labor-intensive, approach. Two findings in our study bolster confidence that we do profile RNA originating from the patient’s cancer cells. First, PCA3, a marker with known prostate cancer specificity, is differentially abundant in this cohort. Second, the TMPRSS2:ERG fusion transcript, known for its prostate cancer specificity, was detected in one of the samples obtained from a prostate cancer patient. As the TMPRSS2-ERG fusion transcript has only been detected in prostate cancer cells and it is not transcribed in normal cells, the detection of TMPRSS2-ERG supports the presence of tumor-derived mRNA in seminal plasma.

Twelve high-confidence fusion transcripts were identified in our cohort. Only 2/12 transcripts were previously detected in prostate cancer tissue. Even though stringent filtering steps were applied to avoid false positive fusion transcripts, further validation of the presence of the fusion transcripts in seminal plasma using RT-qPCR is necessary. Fusion transcript BMS1-PTPN20 has been identified in two cancer samples and one healthy individual, suggesting that its contribution to the pathogenesis of prostate cancer might be limited. The TMPRSS2:ERG fusion transcript results in an increased expression of the ERG oncogene and has been identified in half of the prostate cancer tissues as driver of the disease^24^. The TMPRSS2-ERG fusion transcript has also been investigated in urine as potential liquid biopsy component for the detection of prostate cancer. Tomlins et al. reported the development of the Mi-Prostate Score, an algorithm consisting of the serum PSA level combined with urine TMPRSS2:ERG fusion transcript abundance and urine PCA3 mRNA abundance to predict the risk of detecting prostate cancer on a tissue biopsy^32^. McKiernan et al. developed a urine-based gene expression assay that discriminates high-grade from low-grade prostate cancer and benign disease in order to reduce the number of unnecessary prostate tissue biopsies^33^. While both assays seem promising, their clinical utility remains to be determined.

Two important limitations of seminal plasma as a source of biomarkers are the potential difficulty in obtaining semen from elderly men with erectile dysfunction and the potential personal objections to donate this fluid owing to ethical or religious considerations^34,35^.

Biomarkers that can accurately detect prostate cancer at an early stage and identify aggressive disease subtypes are needed to improve patient management. In this study we provide proof-of-principle that extracellular RNAs in seminal plasma can be profiled and contain -amongst others- mRNAs that are derived from tumor cells. Further investigation in a larger, prospectively collected cohort using standardized collection procedures and optimized RNA purification is warranted. These future cohorts should also contain a higher proportion of samples obtained from patients with aggressive disease (Gleason > 8), as this group is underrepresented in our study.

## Data Availability

The raw RNA-sequencing data is currently being deposited at the European Genome-phenome Archive (EGA).

## Data availability

The raw RNA-sequencing data will be deposited at the European Genome-phenome Archive (EGA).

## Code availability

The Python and R scripts to reproduce the analyses and plots reported in this paper are available from the corresponding authors upon request.

## Contributions

J.V. and P.M. conceived and supervised the project; E.H. and J.N. designed and performed the experiments; E.H., A.M. and L.H. analyzed the data; E.H., G.P., M.M., S.L., E.D., J.K., I.P., and N.L. collected samples; E.H. drafted the paper; All authors contributed to manuscript editing and approved the final draft.

## Supplemental figures

**Supplemental figure 1.**
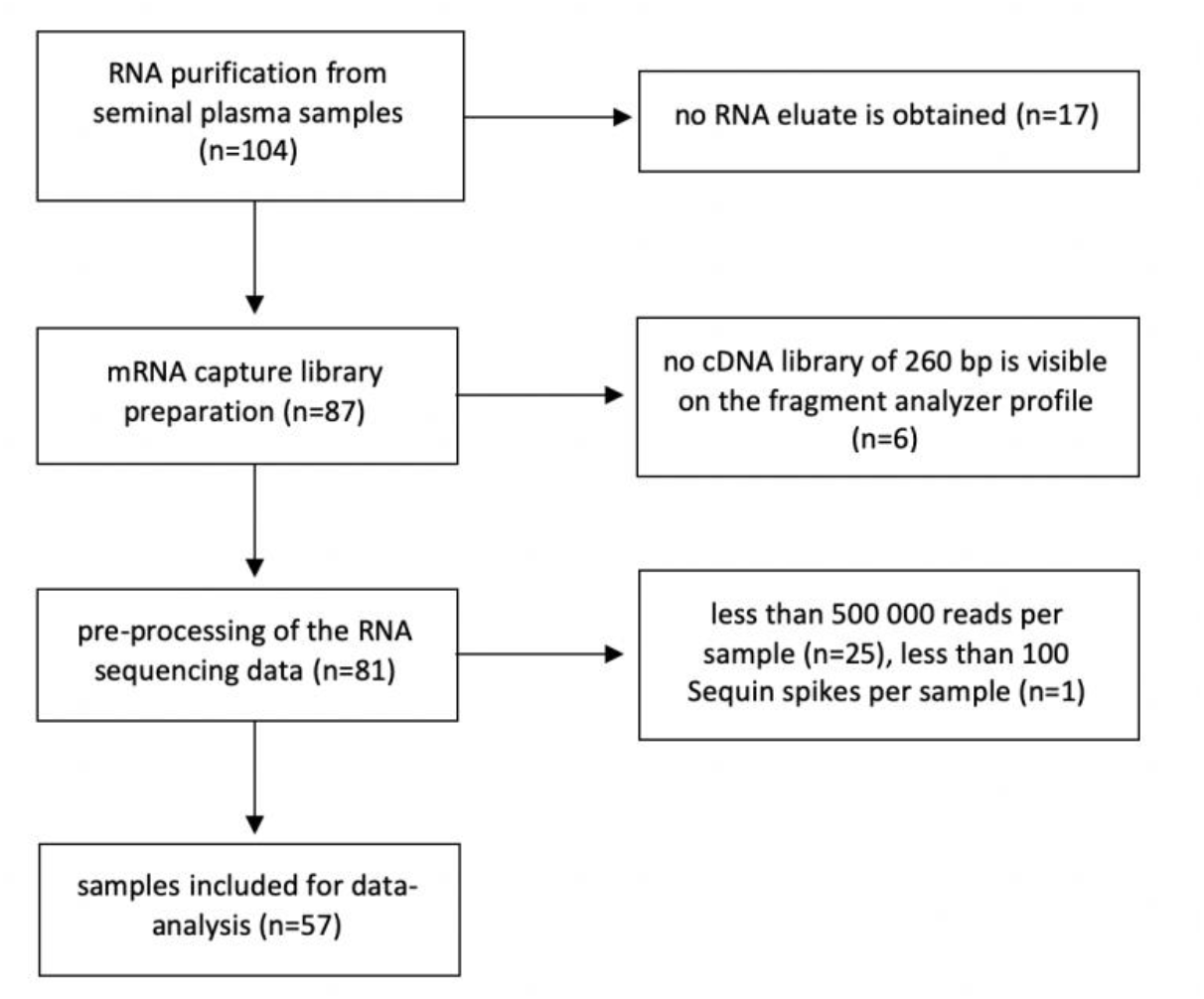
Flowchart justifying the exclusion of seminal plasma samples from the final data-analysis

**Supplemental figure 2.**
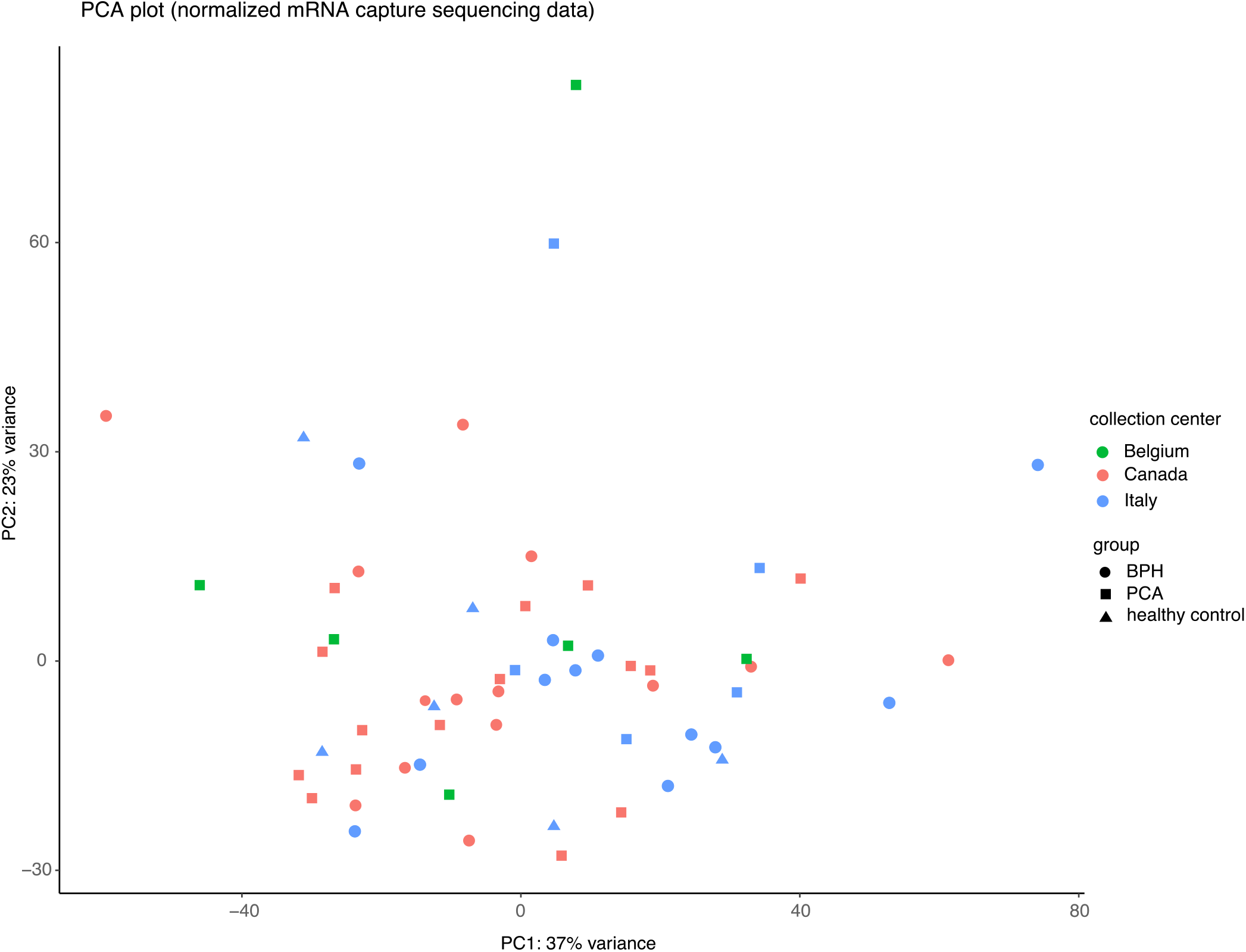
Principal component analysis performed on the normalized sequencing data of all genes across all 57 samples. PCA, prostate cancer; BPH, benign prostate hyperplasia

## Supplemental tables

Supplemental table 1. An overview of the Sequin and ERCC spike-in controls with their stock concentration.

Supplemental table 2. Demographics and clinical characteristics of the patients. Per sample the number of total reads sequenced (totalreads_fastqfile), the total reads after quality control and after adapter trimming (totalreads_afterQC_afteradaptertrimming) and the total reads after PCR duplicate removal (totalreads_afterdeduplication) is provided.

Supplemental table 3. Sequin normalized count table.

Supplemental table 4. Output of the differential expression analysis using DESeq2, comparing prostate cancer samples versus samples of patients with benign prostate hyperplasia and samples of healthy individuals.

Supplemental table 5. Fusion description labels indicated in red have a high probability of being false positive and are filtered out for further analysis. High confidence fusion transcripts are defined as fusion transcripts with only orange or green fusion description labels. High confidence fusion transcripts detected in seminal plasma samples are listed. Per fusion transcript the fusion partners with their genomic location, the fusion description label and the predicted effect of the gene fusion are provided.

